# Engaging patient communities in intracranial neuroscience research

**DOI:** 10.64898/2026.04.14.26350320

**Authors:** Ashley Walton, Erika Versalovic, Amanda R. Merner, Gabriel Lázaro-Muñoz, Alan Bush, R. Mark Richardson

## Abstract

Patients who participate in intracranial neuroscience research make invaluable contributions to our understanding of the brain, accelerating the development of neurotechnological interventions. Engagement of patients as part of this research presents unique challenges, where study goals can be distant from immediate clinical applications and require specialized domain knowledge. Yet methods for meaningfully integrating patient communities as part of these research efforts is essential, as intracranial neuroscience guides the application of artificial intelligence for understanding and enhancing human cognition. In order to identify what patients consider meaningful research engagement we interviewed individuals who participated in a study during their Deep Brain Stimulation (DBS) surgery and attended a group event where they interacted with our research team. Analysis of semi-structured interviews identified four main themes: interest in science and the future of clinical care, contributing to science to improve lives, connecting with others, and accessibility considerations. Based on these insights, we propose strategies for transformational participation of patient communities in intracranial neuroscience research with respect to engagement objectives, communication and scope. This approach offers a foundation for sustaining relationships between scientists and communities rooted in trust and transparency, to ensure that impacts of neurotechnology on human health and cognition are aligned with patient needs as well as desired public values.

## INTRODUCTION

Patients implanted with Deep Brain Stimulation (DBS) who participate in intracranial neuroscience studies contribute foundational knowledge to our understanding of brain function, how this function is impaired by disease, and how neurostimulation mitigates disease symptoms. As opportunities to record intracranial data are rare, patients undergoing neurosurgical therapies are uniquely positioned to contribute to advances in human neuroscience. Brain activity recorded from their implanted devices provide an indispensable contribution to our understanding of neural function and is necessary for designing more effective treatments.

In order to develop treatments that support a wide range of patient needs, research studies must engage multiple individuals with different disease experiences to build a comprehensive understanding of the brain and behavior. Participatory research methods can support active engagement between patients and research teams [1,2], where there are frameworks for structuring patient partnerships in healthcare research [3] as well as medical device development [4]. But there is less guidance for how to structure their inclusion as part of basic studies in neuroscience that are foundational to the development of these devices. And there are unique challenges to engaging patients in these projects where the primary objective is to understand brain function. First, while patients are often motivated to contribute for the benefit of individuals with their condition [5], the implications of study findings on clinical care are still unknown. Researchers must balance communicating study objectives in accessible language while being mindful of over-promising future translational value—considerations necessary to avoiding therapeutic misconception or issues of informed consent [6]. Second, when a research study is focused on developing a clinical treatment, patients are experts on the lived experience of the disease and can make direct, explicit contributions to study goals (e.g., provide guidance on barriers and facilitators to participation, help identify research questions based on their experience with the condition). However, basic studies in neuroscience test pre-defined research hypotheses driven by domain knowledge, limiting the scope of patients’ involvement in more foundational stages of projects.

Despite these challenges, intracranial neuroscience particularly needs strategies for integrating participants into the scientific process—both for the benefit of patients and research efforts more broadly. Advances in artificial intelligence continue to increase the potential for neurotechnological devices to significantly transform human cognition and behavior; relationships between scientists and patient communities are essential to ensuring these transformations contribute to desired societal outcomes [7,8]. Patient involvement in basic science studies has been shown to improve research quality and efficiency, but their contributions are limited to more passive roles [9-11] as this research requires specialized expertise and their impact on clinical care progresses at the timescale of years or decades. This creates barriers to scientists understanding how their efforts ultimately impact the lives of those who benefit from their work and prevents patients from appreciating the significance of their participation. Meaningful engagement in these studies doesn’t require patients to take on the responsibilities of researchers [12], but it does necessitate equitable relationships that are mutually beneficial to both parties [10,11,13,7].

But what do patients find meaningful about engaging with intracranial neuroscience research? What kinds of interactions add value to their research participation? To answer this question, we employed qualitative research methods to understand patients’ interests in contributing to neuroscience research and what they found meaningful about an event designed to engage them with our research team. We summarize interview insights and translate our findings into strategies for engaging patient communities from what we learned about their motivations and interests. We conclude with a discussion of how this approach can provide initial groundwork for supporting exchanges between the field of neuroscience and the public more broadly.

### Intraoperative neuroscience study investigating neural mechanisms of speech

The Brain Modulation Lab at Massachusetts General Hospital (MGH) conducts intracranial neuroscience research; for this current study we interviewed patients who participated in a speech task as part of the implantation of DBS. Brain activity is recorded from where their DBS leads are implanted during their surgery (either subthalamic nucleus (STN), globus pallidus interna (GPi), globus pallidus externa (GPe) or the ventral intermediate nucleus of the thalamus (VIM)), and from electrodes temporarily placed on the surface of their brain over regions that contribute to speech production and perception (see *Figure 1*). The purpose is to understand how different parts of the brain communicate with each other to produce speech, where DBS implantation provides the only clinical scenario for recording neural activity directly from these subcortical and cortical regions of the human brain at the same time. This research has the potential to inform future approaches to developing DBS for addressing speech symptoms in movement disorders.

**Figure 1.**
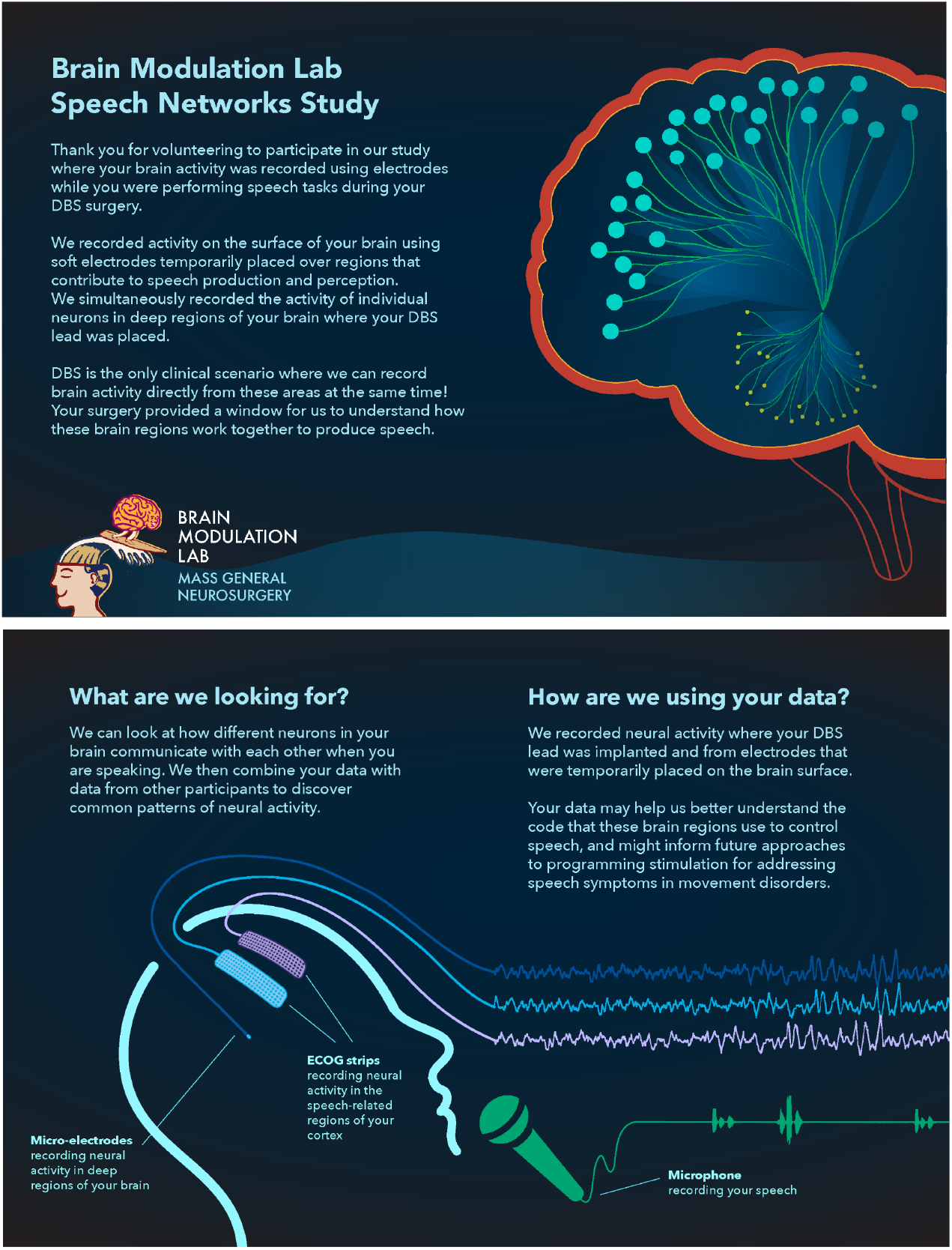
Schematic of intraoperative speech task from study brochure. To improve communication about study information author A.W. designed a brochure that included visuals, description of study goals and information about how their data would be used.

## METHODS

### Research Event

We designed an event in collaboration with a patient-partner to share preliminary findings from the intraoperative speech study and further engage participants with our neuroscience research team. Twenty patients who participated in the study were contacted by phone and invited to attend the research event in October 2024. Twelve patients and eight caregivers attended the two and a half hour event held at Massachusetts General Brigham’s Assembly Row campus (for demographics, see *Table 1*). It brought together groups that contribute to research related to DBS with content focused on neuroscience, neuroethics, and a group discussion developed by our patient-partner related to DBS and clinical care. There was an hour and a half of presentations: the first by principal investigator and neurosurgeon about the history of DBS (author R.M.R), a presentation of preliminary research findings by faculty instructor (author A.B.), and a presentation by the MGH Brain Bioethics group focused on public perceptions of different neurointerventions (authors A.M., E.V. & G.L.) [14]. Preceding the event, attendees were emailed a draft of a guidebook written by author R.M.R. for patients considering DBS surgery. The last hour of the event was a discussion session led by our patient-partner about the guidebook and what resources patients found valuable before and after DBS surgery (for discussion questions see *Appendix A*). Patients were also given images of their brain activity recorded as part of the research study printed on acrylic blocks (see *Figure 2*). Breakfast pastries and coffee were provided, and patients’ parking was covered for the day.

**Table 1:** Participant demographics.

|  |
| --- |
| **Gender:** Female **7** (58.3%), Male **5** (41.6%) |
| **Race and Ethnicity:** Black **1** (8.33%), Hispanic or Latino/a/e **1** (8.33%), White **10** (83.33%) |
| **Age:** Mean **68**, Min-Max **55-79** |
| **Condition:** Dystonia **1** (8.33%), Essential Tremor **3** (25%), Parkinson's Disease **8** (66.67%) |
| **DBS Target:** GPi **3** (25%), STN **6** (50%), VIM **3** (25%) |
| **Total household income (before taxes) from all sources in the last year:** \$0 to \$49,999 **2** (16.67%), \$50,000 to \$99,999 **4** (33.33%), \$100,000 to \$149,999 **3** (25%), \$150,000 or more **3** (25%) |
| **Source(s) of health insurance:** Employer **2**, Medicare **6**, Private health insurance **9**, State Insurance **2** |

**Figure 2.**
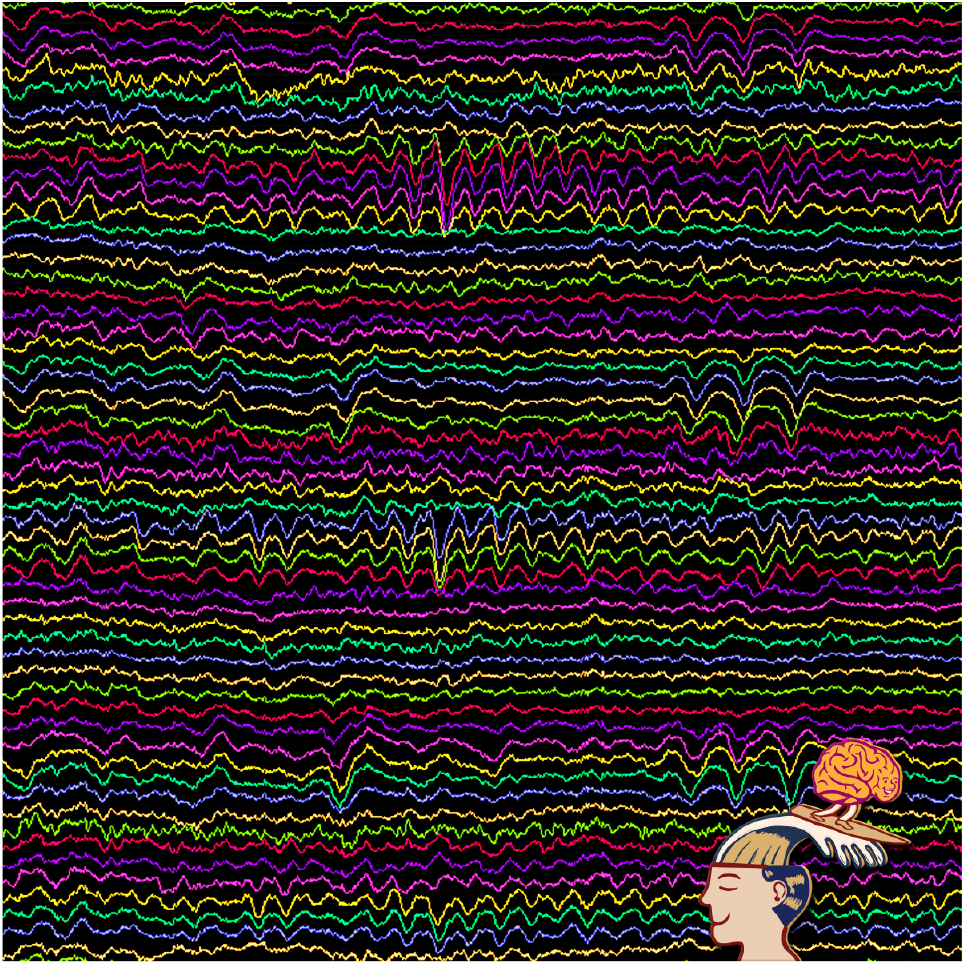
Patient’s brain data recorded as part of intraoperative study, provided to participants who attended the research event.

### Post-Research Event Interviews

We conducted follow-up interviews with attendees to understand what they found meaningful about the event and their interests in being part of neuroscience research. Semi-structured interviews were conducted using video conferencing software (HIPAA-compliant Zoom) and patients were compensated with a $50 gift card for their time. One patient could not attend the event due to an illness but still participated in the interview. Questions focused on what patients found meaningful from the research event and their interests in contributing to neuroscience research (see *Table 2*). These questions were part of longer interviews that aimed to understand patients’ needs after DBS implantation, which in total lasted between 45-90 minutes.

**Table 2:**
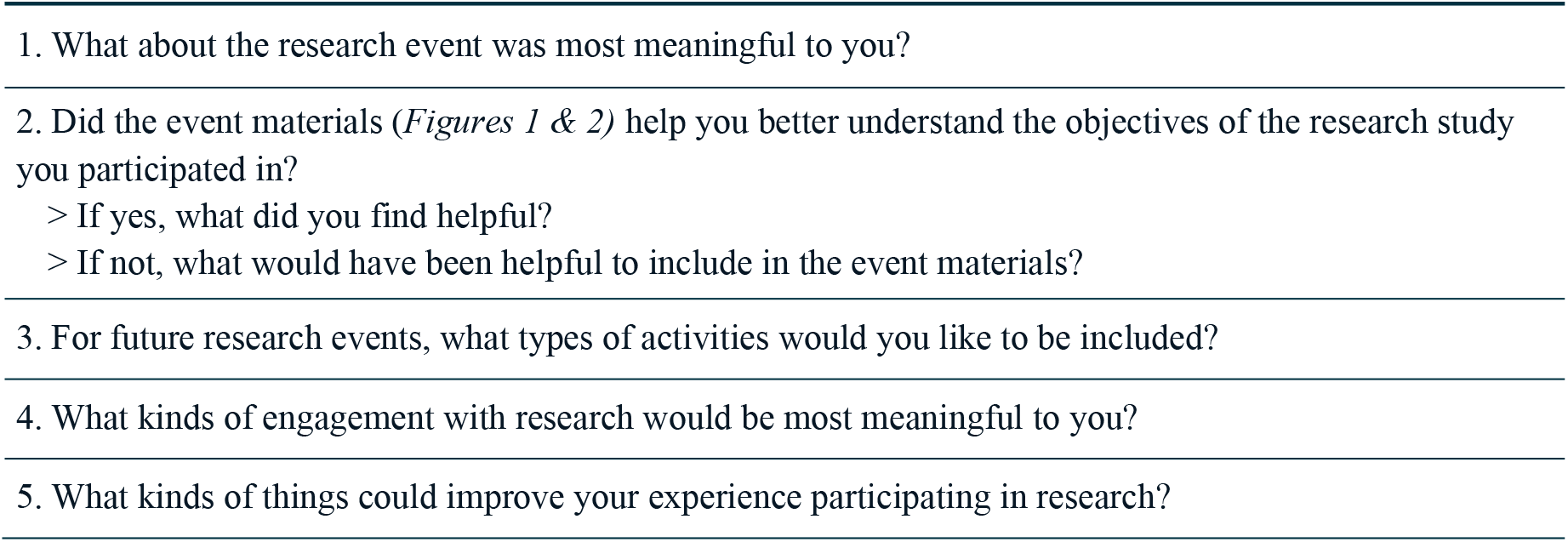
Research event interview questions.

## ANALYSIS

Interview recordings were transcribed, de-identified, and then analyzed using Dedoose Version 10 [15]. Three members of the research team were involved in the qualitative data analysis: one trained in philosophy (author E.V.) and two experimental psychologists (authors A.W. & A.M.), all with extensive backgrounds in neuroethics and DBS research. Thematic content analysis was used to iteratively identify themes in the data [16], where two team members coded each individual transcript and the third team member reviewed the codes and led discussions to define consensus in code definition and interpretation.

## RESULTS

Interview analysis resulted in four main themes: Interest in science and the future of clinical care, Contributing to science to improve lives, Connecting with others, and Accessibility considerations (for subthemes see *Appendix B*).

### 1. INTEREST IN SCIENCE & THE FUTURE OF CLINICAL CARE

Patients expressed interest in the scientific content (7/12), where two patients wanted to know more study results (see *Table 3*, P8-A & P9), one patient asked for analysis details (see *Table 3*, P4), and two desired to know more about future research trajectories related to clinical care (see *Table 3*, P11 & P8-B).

**Table 3:** Participant descriptions of their interest in science and the future of clinical care.

|  |
| --- |
| **Q: What kinds of things could improve your experience participating in research?** |
| ● *I've been in a number of research studies and this one was the only one where I got to hear feedback, or anything about it afterwards, which I found very helpful and refreshing...Because the other studies, you're just like, oh, well, what happened with all that?* (P8-A) |
| **Q: What kinds of engagement with research would be most meaningful to you?** |
| ● *I'd just like to know the results. I participated in a spinning study when I first got diagnosed with Parkinson's years ago. And I've never heard the results from that study.* (P9) |
| **Q: What would have been helpful to include in the event materials?** |
| ● *What do you do with the data afterwards? How is it processed? That would be interesting.* (P4) |
| **Q: For future research events, what types of activities would you like to be included?** |
| ● *It might be helpful to know what the team's interested in and how the discussion at the end of the event connects to what the future would be in terms of research. I'm fine to be told "I don't know, we're not sure yet". That's okay, but it'd be nice to know what the vision is— where you think you're going with it.* (P11)<br>● *I think the future of DBS. It might be interesting for me to know the ways those things are coming together.* (P8-B) |

### 2. CONTRIBUTING TO SCIENCE TO IMPROVE LIVES

Participants’ responses to interview questions about their interest in scientific research demonstrated their priority to improve clinical care for others (10/12). Two patients described feelings of pride from better understanding their contributions (see *Table 4*, P8 & P11-A). For two patients, their interest in the neuroscience materials was dependent on its relevance to their clinical condition, specifically whether they experienced issues with their speech (see *Table 4*, P2 & P11-B).

**Table 4:**
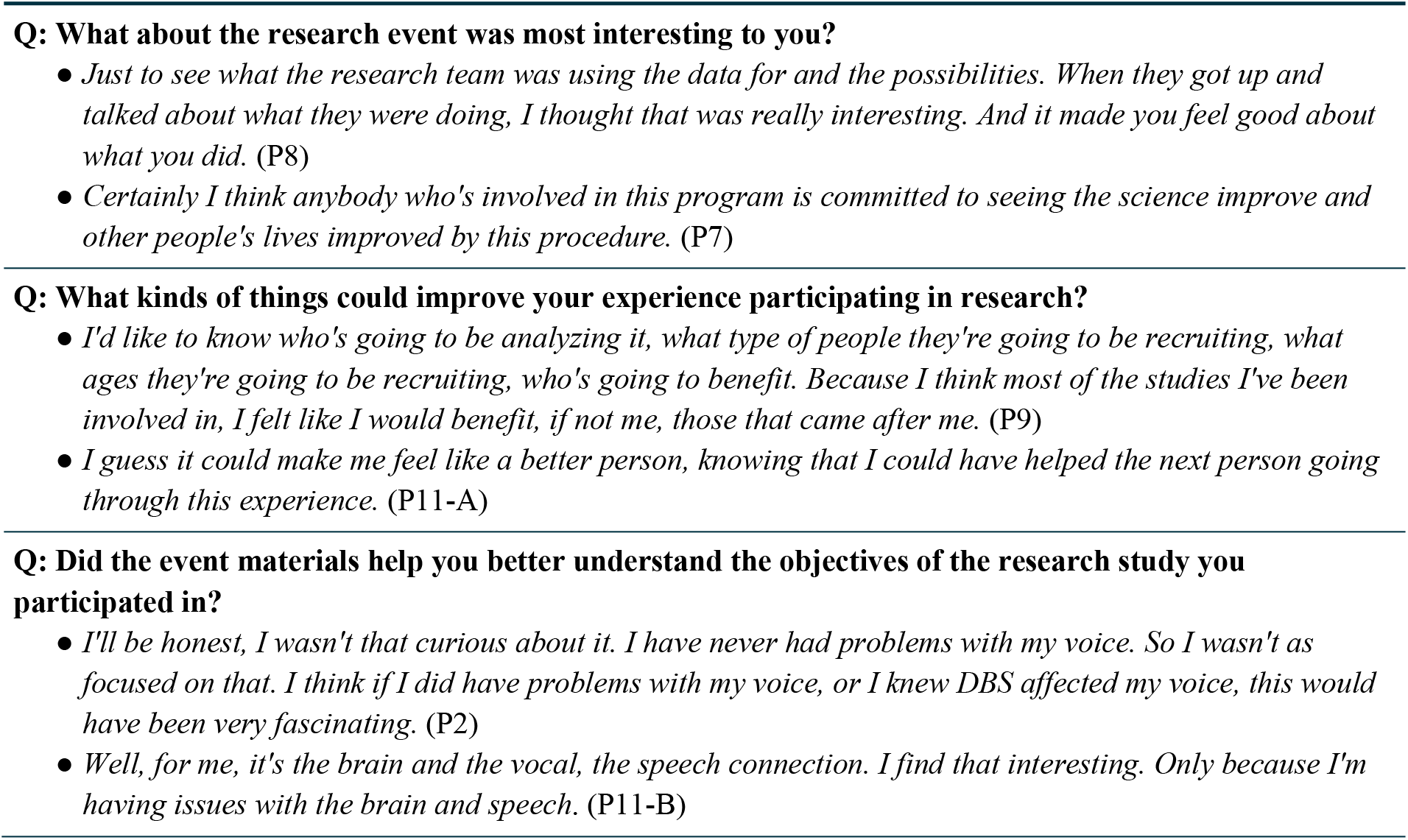
Participant descriptions of contributing to science to improve lives.

### 3. CONNECTING WITH OTHERS

All patients who attended the event identified the opportunity to talk to other patients with DBS as a meaningful aspect of their participation (11/11). The majority of participants (7/11) specifically mentioned the value of pooling knowledge, and hearing about others’ experiences with managing symptoms and DBS treatment. Notably, two individuals that attended the same recreational center with programming for individuals with movement disorders were less focused on this component of the research event. They did not report the same need for social support and felt fortunate to have access to that resource (see *Table 5*, P5-A). Many participants also identified the opportunity to connect with the research team as meaningful (5/11).

**Table 5.**
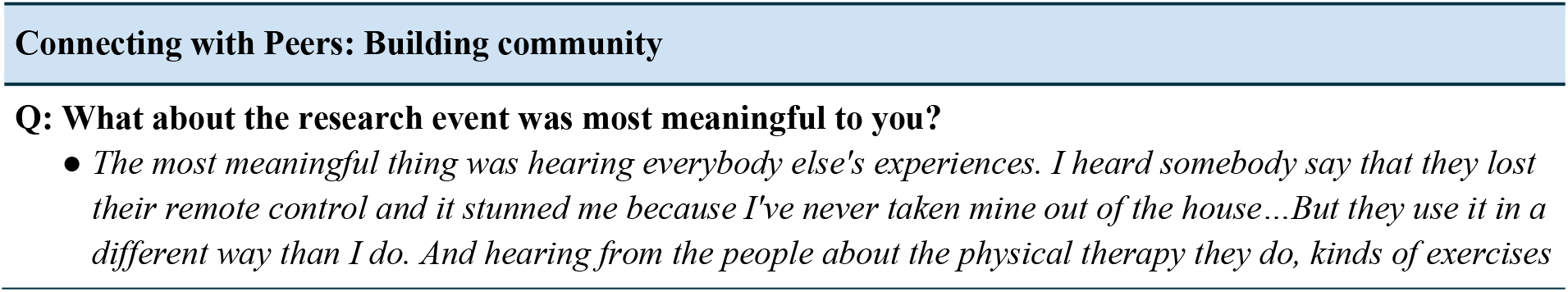
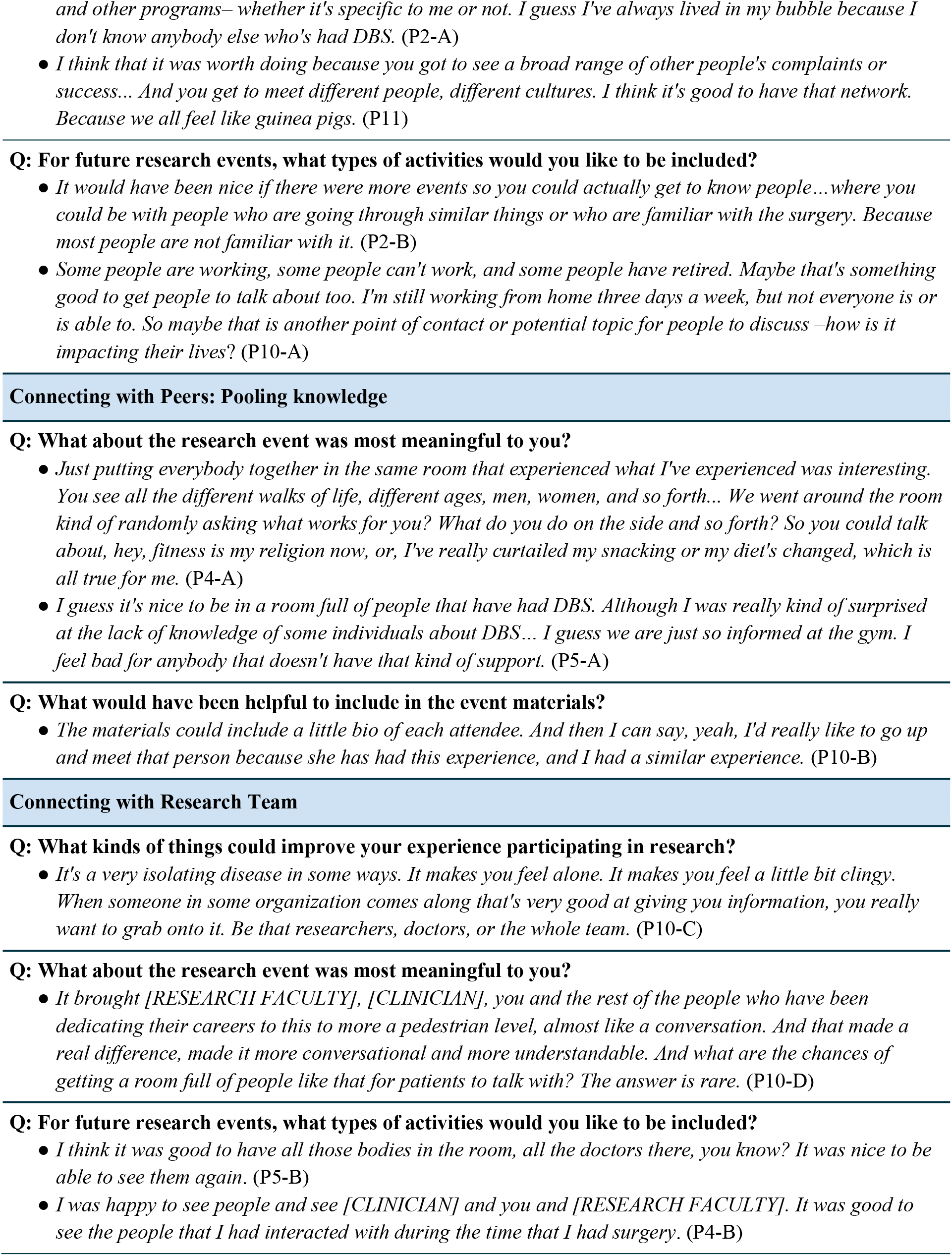
Participant descriptions of connecting with others.

### 4. ACCESSIBILITY CONSIDERATIONS

Patients reported cognitive and communication barriers that impacted their ability to participate in the group discussion at the research event (3/12). For the event materials, one patient described how visuals were helpful, and another requested that we provide a list of event attendees to help them introduce themselves and serve as a reference after the event (see *Table 6*, P10 & P2). One patient described feeling unable to participate in the group discussion and became distressed during the interview when trying to recall the event to answer our questions (see *Table 6*, P12-B).

**Table 6:**
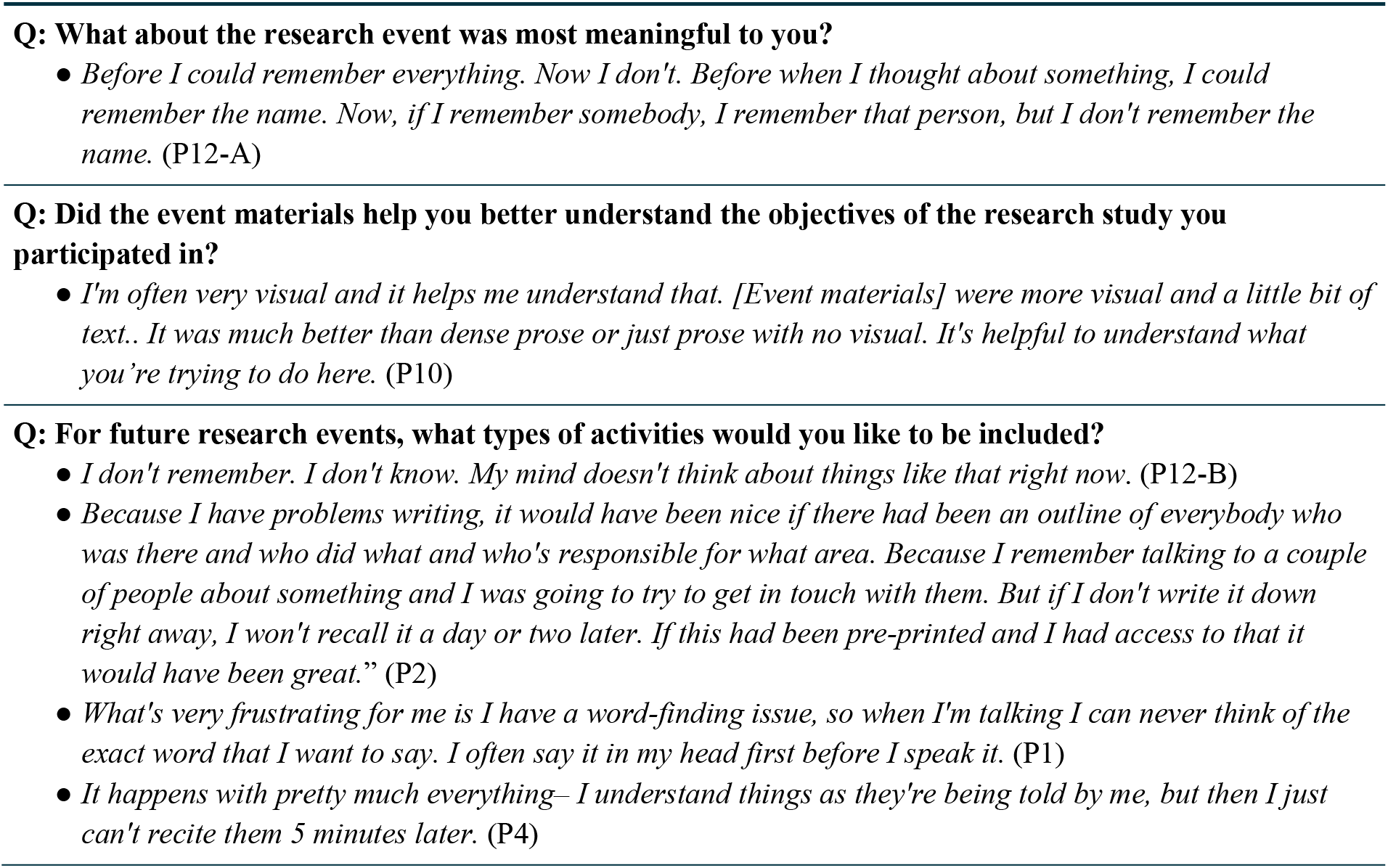
Participant descriptions of accessibility considerations.

## DISCUSSION

### Interest in science and future of clinical care

Patients’ motivation for adding a research study to their surgical procedure was to advance science for improving the lives of others with the same condition. Their goal was to contribute knowledge about the brain, and by extension, knowledge about their disease that might improve clinical care. From the patients’ perspective, they are undergoing surgery because they have a brain disorder, and so they expect recordings of their brain activity to contain information about their disease. This is not a misconception— while the research design is focused on characterizing healthy brain function, doing so requires identifying and distinguishing pathological neurophysiology. During consent the research team focuses on the study’s scientific goals to avoid misleading patients about potential personal benefits. But the project includes the possibility of investigating how features of patients’ brain recordings relate to their pre- and post-implantation symptoms, which could inform treatment development. Focusing only on the immediate scientific goals may unintentionally obscure patients’ understanding of potential future impacts of their participation—something our participants identified as deeply meaningful. Engaging patients in conversations about the possible long-term impact of their research participation, even if outside the study’s scope, is better aligned with their motivations and can provide a foundation for longer-term, collaborative partnerships.

### Contributing to science to improve lives

Patients’ focus on clinical outcomes should be expected, as they are actively managing a disease and understand first-hand the value of contributing toward easing this burden for others. Researchers in neuroscience may have similar altruistic motivations to improve patient lives, though their immediate focus is ensuring their study produces meaningful results. Similarly, patients have been found to express conditional altruism, where initial interest in research participation is driven by helping others, but enrollment is contingent on relevance to their clinical condition and absence of personal harm [5,17] (see *Table 4*, P2, P11-B). But while both patients and researchers have individual concerns that could reasonably take precedence over their altruistic motivations, it is essential for engagement practices to ensure that the burdens and benefits of the research process are equitably distributed [18].

### Connecting with others

There was no single mode of engagement preferred by all patients (see subthemes in *Appendix B*). Some desired more active and participatory forms of engagement such as a patient council or DBS community groups, others reported that a personal phone call or newsletter with updates would feel meaningful. But while our findings suggest there is no single mode of engagement that participants prefer, we found that all patients had similar goals that were primarily social and prosocial in nature. They all highly valued the opportunity to connect with each other and learn from each other’s disease experiences. Facilitating connection around these shared experiences is critical for meaningful engagement of this patient population in neuroscience research.

### Accessibility Considerations

Patients’ contributions are often contingent on disease burden, which varies depending on financial resources, care support, symptom severity and disease progression. Our interviews identified some accessibility considerations that could support patients in equally participating in research engagement. For example, challenges they reported with writing and recall can inform the materials made for future research events (see *Table 6*) where we could provide a list of participant bios with accompanying photographs. Additionally, making overviews of research event topics available prior to the event could help those who need more time to cognitively process and reflect on what kinds of questions they may want to ask. For patients that experiencing memory impairments (see *Table 6*, P12-A, P12-B), we could provide an overview of topics that will be covered prior to the interview, add more illustrative descriptions of aspects of the events being referenced in our questions, and include their caregiver using a dyadic interview method (for example see [19]). Research engagement can include general tools that support the active involvement of all patients, while some strategies must be tailored to patients’ specific challenges, and responsive to changes in their needs over time.

## PARTICIPATORY STRATEGIES FOR ENGAGING PATIENT COMMUNITIES IN NEUROSCIENCE

In joining efforts towards a more translational neuroethics [18], we build upon our interview insights and experience working with our patient-partner to propose strategies for implementing participatory engagement as part of intracranial neuroscience research. Our approach adds to tools developed for engaging patients in laboratory research [20,21] and builds upon the concepts outlined by Pandaya et al. (2025) regarding transactional versus transformational engagement [7]. They describe transactional engagement as short-term, limited patient involvement focused on an individual problem. Transformational engagement entails long-term relationships for collaboratively identifying and addressing needs of both scientific and participant communities. We provide strategies for how three main components of research engagement: objectives, communication and scope, can be made transformational so that neuroscientific discovery systematically includes all who contribute to its progress.

### 1. Engagement objectives: Prioritize a patient-valued goal

Our patient-partner discussed the need for resources to support patients before and after surgery. He wanted to understand their experiences with DBS and identify what information they would find valuable. So in collaboration with author A.W., he created a webpage for the lab’s website that includes resources for prospective patients and the DBS guidebook edited to incorporate feedback we received during the group discussion at the research event. In this case participatory engagement was not focused on a shared scientific goal, instead study goals were pursued in parallel while working with our partner to make more clinical resources available to the patient community. The use of parallel outcomes can ensure that relationships between scientists and communities are mutually beneficial [7] and obviates challenges to identifying relevant intersections between the study’s scientific goals and patients’ interests or expertise. While they themselves won’t directly benefit from their engagement with basic studies in neuroscience, pursuing a patient-valued goal allowed our partner to improve the lives of others in the short-term (making more clinical resources available to patients) while the research team’s scientific efforts contribute towards impact in the longer term (identifying the neural mechanisms of speech). Transformational neuroscience engagement requires recognizing the importance of applying research team efforts towards a goal valued by patients even if it does not directly contribute to the scientific objectives of an individual study.

### 2. Engagement Communication: Co-develop materials about the short and long-term research goals

We found that patients desired greater insight into the future of DBS research and its potential impact on clinical care (see *Tables 3 & 4*). We then worked with our patient-partner to clarify and expand the lab’s visual materials focused on future research directions and articulate potential paths towards clinical applications. This provided us an opportunity to better understand gaps that exist between our research studies and their application to clinical practice. For example, when discussing visual materials about the study objectives with patients (shown in *Figure 1*), they reflected on their own challenges with speech and communication. This increased the research team’s awareness of the range of effects DBS patients experience, resulting in a new project focused on assessing speech outcomes after surgery. Transformational neuroscience engagement includes regular communication that continuously aligns research efforts with patient needs and develops a shared understanding of the broader impacts of the team’s collective efforts.

### 3. Engagement scope: Organize regular events to build community

We found that the opportunity to connect with other patients was the most valued component of our research engagement efforts (see *Table 5*), suggesting that this is the most significant way we can make neuroscience research engagement beneficial and meaningful to our patient community. For example, they requested more opportunities for personal exchanges as part of the research event, including small-group sessions and additional time set aside for socializing. Inspired by the event, our patient-partner himself initiated a monthly discussion group for individuals with movement disorders at his local community center. Creating regular opportunities for patients to connect with one another allows them to build community around their research contributions and disease experience. These events can also help sustain long-term relationships between patients and researchers that enable them to effectively collaborate as research objectives evolve [22]. Transformational neuroscience engagement expands the scope of engagement beyond individual grant-funded projects, making longitudinal partnerships foundational to a lab’s research agenda.

## CONCLUSION

We interviewed patients to understand what they found meaningful about research engagement and outlined strategies to build relationships between scientists and patient communities for collaboratively shaping and strengthening neuroscience research efforts. These are defined from what our participants reported as meaningful and may not generalize to groups outside of individuals with movement disorders seeking DBS. The goal of our strategies is to enable other teams to adapt and apply what we found within their own research contexts, offering a starting point for a broader vision of engagement efforts.

The future of this work will consider these partnerships across other diseases and research modalities (e.g., intracranial monitoring in the epilepsy monitoring unit), as well as across institutions, especially for neuroscience studies where combining datasets across centers is essential to generalizable insights that can more significantly impact clinical care. New initiatives focused on data standardization, reproducibility and database development [23-26] are aligned with patient priorities to increase the impact of their participation by ensuring their neural recordings can be used across institutions to accelerate research progress [27]. This presents new opportunities for patients to appreciate the additional contributions of their data, as well as build awareness around data sharing practices. Patients currently have limited knowledge regarding how their neural data is included in these repositories and have no role in shaping how it is used for public or private development. A new challenge is engaging patient communities as part of secondary analyses projects, where there are currently no touchpoints for integrating participant input.

We see these strategies as a step towards extending the goals of engagement beyond both individual and multi-center studies to support a larger infrastructure for aligning neuroscientific efforts with public values. Systematic approaches for maintaining long-term relationships between scientists and research participants are necessary to a collaborative process for guiding neurotechnological advances [8]. Publications communicate scientific results *to* the public, but resources are needed for sustaining communication about the ambitions of science *with* the public. As one of our participants said: “*I’m fine to be told ‘I don’t know, we’re not sure yet’. That’s okay, but it’d be nice to know what the vision is– where you think you’re going with it*.” Engagement strategies outlined here can build relationships that allow scientists and patient communities to aggregate what they know–as well as also work together with transparency about what they don’t know–towards the common goal of advancing science and its ability to shape and expand human cognition.

## Acknowledgements

The authors thank our patient-partner Thomas Jones who collaborated with us to create the research event and continues to donate his time and insight to our patient engagement team. This work was supported by grants from the National Institute of Neurological Disorders and Stroke, Award Number 5U01NS117836-05 (P.I. R. Richardson), and the National Institute of Mental Health, Award Number 5R01MH133657-02 (P.I. G. Lázaro-Muñoz).

## Statements and Declarations

### Ethics Approval

This study was performed in line with the principles of the Declaration of Helsinki. Approval was granted by the Mass General Brigham Institutional Review Board (Protocol #2022P000223).

### Consent to Participate

Informed consent was obtained from all individual participants included in the study.

### Consent to Publish

Not applicable.

### Data Availability

The datasets generated during the current study are available from the corresponding author on reasonable request.

### Author Contributions

All authors contributed to conceptualization of the project. A.W. wrote the original draft and all others authors reviewed and edited. A.W. and E.K. conducted data collection, A.W., E.K. and A.M. data analysis. G.L. and R.M.R. provided funding and supervision. A.W. created all figures.

## APPENDIX A

*Questions for the discussion session led by our patient partner at the research event*.

### Questions about the DBS guidebook

- Do you have any thoughts/feedback on the guidebook?
- What parts of the book did you think would have been particularly helpful?
- Are there things that were missing?

### Questions about resources pre- and post-DBS

- How did you learn about DBS, what made you decide to do it?
- What resources have you found for pre-and post-op needs?
- It can be a very isolating disease and each one of us, especially pre-DBS, probably had quite different experiences. Are they any you would like to share?
- What are we doing to help manage our PD?
- What do you do to stimulate your brain?
- What resource do you wish was out there?
- How has DBS changed your lives (patients + caregivers)?

## APPENDIX B

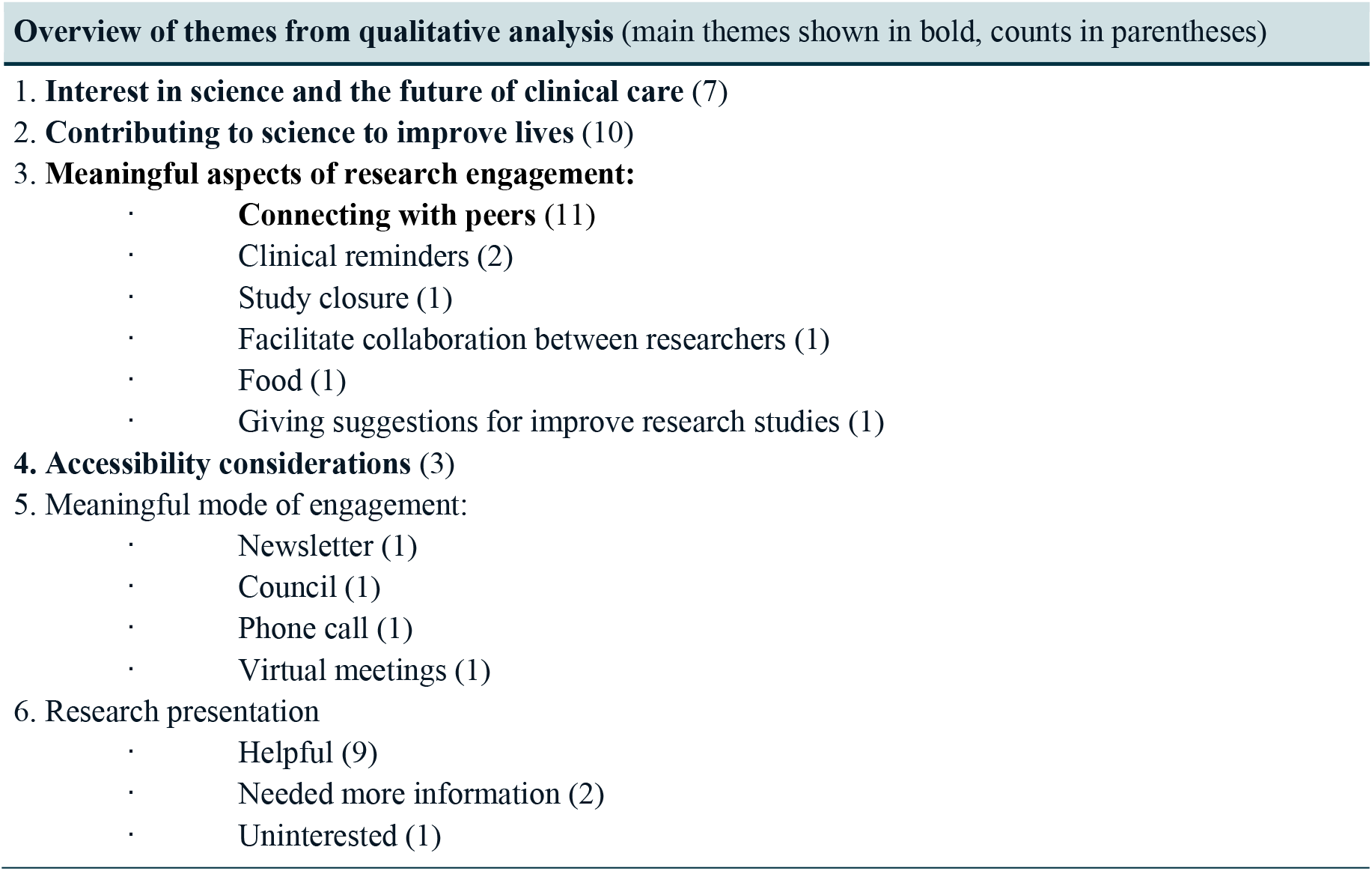

## REFERENCES

1. Montreuil, M., Martineau, J. T., & Racine, E. (2019). Exploring ethical issues related to patient engagement in healthcare: Patient, clinician and researcher’s perspectives. Journal of Bioethical Inquiry, 16(2), 237–248. 10.1007/s11673-019-09904-6

2. Racine, E. (2024). What participatory research and methods bring to ethics: Insights from pragmatism, social science, and psychology. Kennedy Institute of Ethics Journal, 34(1), 99–134. 10.1353/ken.2024.a943431

3. McCarron, T. L., Clement, F., Rasiah, J., Moran, C., Moffat, K., Gonzalez, A., Wasylak, T., & Santana, M. (2021). Patients as partners in health research: A scoping review. Health Expectations, 24(4), 1378–1390. 10.1111/hex.13272

4. U.S. Food and Drug Administration (FDA). (2022). Patient engagement in the design and conduct of medical device clinical studies. https://www.fda.gov/regulatory-information/search-fda-guidance-documents/patient-engagement-design-and-conduct-medical-device-clinical-studies

5. Outram, S., Muñoz, K. A., Kostick-Quenet, K., Sanchez, C. E., Kalwani, L., Lavingia, R., Torgerson, L., Sierra-Mercado, D., Robinson, J. O., Pereira, S., Koenig, B. A., Starr, P. A., Gunduz, A., Foote, K. D., Okun, M. S., Goodman, W. K., McGuire, A. L., Zuk, P., & Lázaro-Muñoz, G. (2021). Patient, caregiver, and decliner perspectives on whether to enroll in adaptive deep brain stimulation research. Frontiers in Neuroscience, 15, 734182. 10.3389/fnins.2021.734182

6. Mergenthaler, J. V., Chiong, W., Dohan, D., Feler, J., Lechner, C. R., Starr, P. A., & Arias, J. J. (2021). A qualitative analysis of ethical perspectives on recruitment and consent for human intracranial electrophysiology studies. AJOB Neuroscience, 12(1), 57–67. 10.1080/21507740.2020.1866098

7. Pandya, R. E., Boyd, A. D., Feliú-Mójer, M. I., & Yanovitzky, I. (2025). Transformative community-engaged science: Strengthening relationships between science and society. Proceedings of the National Academy of Sciences, 122(27), e2400929122. 10.1073/pnas.2400929122

8. Kaebnick, G. E. (2024). Neuroscience and society: Supporting and unsettling public engagement. Hastings Center Report, 54(1), 20–23. 10.1002/hast.1565

9. Fox, G., Fergusson, D. A., Daham, Z., Youssef, M., Foster, M., Poole, E., Sharif, A., Richards, D. P., Hendrick, K., Mendelson, A. A., Macala, K. F., Monfaredi, Z., Montroy, J., Fiest, K. M., Presseau, J., & Lalu, M. M. (2021). Patient engagement in preclinical laboratory research: A scoping review. EBioMedicine, 70, 103484. 10.1016/j.ebiom.2021.103484

10. de Wit, M. P., Koenders, M. I., Neijland, Y., van den Hoogen, F. H. J., van der Kraan, P. M., van de Loo, F. A. J., & van den Ende, C. (2022). Patient involvement in basic rheumatology research at Nijmegen: A three year’s responsive evaluation of added value, pitfalls and conditions for success. BMC Rheumatology, 6(1), 66. 10.1186/s41927-022-00296-6

11. Bird, M., Ouellette, C., Whitmore, C., Li, L., Nair, K., McGillion, M. H., Yost, J., Banfield, L., Campbell, E., & Carroll, S. L. (2020). Preparing for patient partnership: A scoping review of patient partner engagement and evaluation in research. Health Expectations, 23(3), 523–539. 10.1111/hex.13040

12. Spears, P. A. (2021). Meaningful patient engagement in cancer research: How patient advocacy is changing the research landscape. Future Oncology, 17(10), 1173–1178. 10.2217/fon-2020-1198

13. Schölvinck, A. F. M. (2018). Towards meaningful and sustainable patient involvement in health research decision-making [Doctoral dissertation]. Vrije Universiteit Amsterdam.

14. Furrer, R. A., Merner, A. R., Stevens, I., Zuk, P., Williamson, T., Shen, F. X., & Lázaro-Muñoz, G. (2025). Public perceptions of neurotechnologies used to target mood, memory, and motor symptoms. Device, 3, 100804. 10.1016/j.device.2025.100804

15. SocioCultural Research Consultants, LLC. (2025). Dedoose (Version 10) [Computer software]. https://www.dedoose.com

16. Boyatzis, R. E. (1998). Transforming qualitative information: Thematic analysis and code development. Sage.

17. McCann, S. K., Campbell, M. K., & Entwistle, V. A. (2010). Reasons for participating in randomised controlled trials: Conditional altruism and considerations for self. Trials, 11(1), 31. 10.1186/1745-6215-11-31

18. Wexler, A., & Specker Sullivan, L. (2021). Translational neuroethics: A vision for a more integrated, inclusive, and impactful field. AJOB Neuroscience, 12(4), 250–256. 10.1080/21507740.2021.2001078

19. Johansson, I. L., Samuelsson, C., & Müller, N. (2020). Patients’ and communication partners’ experiences of communicative changes in Parkinson’s disease. Disability and Rehabilitation, 42(13), 1835–1843. 10.1080/09638288.2018.1539875

20. Lalu, M. M., Richards, D. P., Foster, M., French, B., Crawley, A. M., Fiest, K. M., Hendrick, K., Macala, K. F., Mendelson, A., Messner, P., Nicholls, S. G., Presseau, J., Séguin, C., Sullivan, P., Thébaud, B., & Fergusson, D. A. (2024). Protocol for co-producing a framework and integrated resource platform for engaging patients in laboratory-based research. Research Involvement and Engagement, 10, 25. 10.1186/s40900-024-00545-7

21. Maccarthy, J., Guerin, S., Wilson, A. G., & Dorris, E. R. (2019). Facilitating public and patient involvement in basic and preclinical health research. PLoS ONE, 14(5), e0216600. 10.1371/journal.pone.0216600

22. Sullivan, M., & Poliakoff, E. (2023). Parkinson’s from inside out: Emerging and unexpected benefits of a long-term partnership. Research for All, 7(1), 1–14. 10.14324/RFA.07.1.01

23. Holdgraf, C., Appelhoff, S., Bickel, S., Bouchard, K., D’Ambrosio, S., David, O., … & Hermes, D. (2019). iEEG-BIDS, extending the Brain Imaging Data Structure specification to human intracranial electrophysiology. Scientific Data, 6(1), 102. 10.1038/s41597-019-0105-7

24. Rübel, O., Tritt, A., Ly, R., Dichter, B. K., Ghosh, S., & Niu, L. (2022). The Neurodata Without Borders ecosystem for neurophysiological data science. eLife, 11, e78362. 10.7554/eLife.78362

25. National Institutes of Health. (2024). Notice of Special Interest (NOSI): Promoting data reuse for health research (NOT-OD-24-096). https://grants.nih.gov/grants/guide/notice-files/NOT-OD-24-096.html

26. Markiewicz, C. J., Gorgolewski, K. J., Feingold, F., Blair, R., Halchenko, Y. O., Miller, E., Hardcastle, N., Wexler, J., Esteban, O., Goncalves, M., Jwa, A., & Poldrack, R. A. (2021). The OpenNeuro resource for sharing of neuroscience data. eLife, 10, e71774. 10.7554/eLife.71774

27. Guerrini, C. J., Robinson, J. O., Crossnohere, N. L., Majumder, M. A., Jones, K. M., Brooks, W. B., Sheth, S. A., & McGuire, A. L. (2025). Privacy in perspective: Research participants’ priorities and concerns related to sharing data generated in human neuroscience studies. Neuroethics, 18, 37. 10.1007/s12152-025-09609-1

